# HUMORAL AND CELLULAR IMMUNOGENICITY and SAFETY UP TO 4 MONTHS AFTER VACCINATION WITH BNT162B2 mRNA COVID-19 VACCINE IN HEART AND LUNG TRANSPLANTED YOUNG ADULTS

**DOI:** 10.1101/2021.09.20.21263836

**Authors:** Nicola Cotugno, Chiara Pighi, Elena Morrocchi, Alessandra Ruggiero, Donato Amodio, Chiara Medri, Luna Colagrossi, Cristina Russo, Silvia Di Cesare, Veronica Santilli, Emma Concetta Manno, Paola Zangari, Carmela Giancotta, Stefania Bernardi, Luciana Nicolosi, Marta Ciofi Degli Atti, Massimiliano Raponi, Salvatore Zaffina, Sara Alfieri, Richard Kirk, Carlo Federico Perno, Paolo Rossi, CONVERS study Team, Antonino Amodeo, Paolo Palma

**Author notes:** **Corresponding author(s):** Paolo Palma, MD, PhD, Clinical & Research Unit of Clinical Immunology and Vaccinology, Academic Department of Pediatrics, IRCCS Bambino Gesù Children’s Hospital, Address: Piazza S. Onofrio, 4- 00165 Rome, Italy, Antonio Amodeo, MD, Departments of Pediatric Cardiology and Cardiac Surgery, IRCCS Bambino Gesù Children’s Hospital, Address: Piazza S. Onofrio, 4- 00165 Rome, Italy. These Authors equally contributed to the present work (Nicola Cotugno, Chiara Pighi, Elena Morrocchi, Alessandra Ruggiero). **AUTHORSHIP** NC, PP and PR participated in research design. NC and PP drafted the first version of the manuscript. All Authors participated to the writing of the final version of the paper. CP, EM, AR, DA, CM, LC, MCDA, MR, SZ, CR, SDC, VS, ECM, PZ, CG, SA, RK, CFP, AA and the CONVERS study team participated in the performance of the research. AA, NC, PR and PP acquired funding. **DISCLOSURE** The authors declare no competing interests.

## Abstract

**Background:** Immunizations among vulnerable population, including solid organ transplant recipients (SOT), present suboptimal responses at vaccination and over time. We investigated safety and immunogenicity of the BNT162B2 mRNA COVID-19 vaccine in 34 SOT young adults as compared to 36 healthy controls (HC).

**Methods:** immunogenicity was measured through the analysis of anti SARS-CoV2 IgG Antibodies and antigen specific CD4 T cells (CD40L+), detected by flow cytometry before vaccination, 21 days after priming (T21), 7 days after booster dose (T28) and 2-4 months after priming (T120). Baseline T and B cell immune phenotype was deeply investigated. The safety profile was investigated by close monitoring and self-reported diary.

**Results:** Anti-S and anti-Trimeric Ab responses were significantly lower in SOT vs HC at T21 (p<0.0001) and at T28 (p<0.0001). Ten out of 34 SOT (29%) at T28 and 3 out of 33 (9%) at T120 had undetectable SARS-CoV-2 IgG. The analysis of SARS-CoV-2 specific CD4 T cells showed lower CD40L expression after *in vitro* stimulation in SOT compared to HC. Lower frequencies of memory B cells were found in patients not responding to vaccination. Lack of seroconversion was higher in patients treated with mycophenolate (p=0.0005). The vaccination was safe and well tolerated. Only short-term adverse events, were reported and no hospitalization or graft rejection were observed after vaccinations.

**Conclusions:** These data show that SOT have a suboptimal immune response following mRNA vaccinations as compared to HC. Alternative strategies should be investigated to improve the immunization against SARS-CoV-2 in these patients.

## INTRODUCTION

Large series of Solid organ transplant recipients (SOT) affected by SARS-CoV-2 infection were recently reported and showed more severe lymphopenia and higher case fatality rate compared to the general population^1–3^. Besides, the quantification of SARS-CoV-2 viral load performed on longitudinally collected nasopharyngeal swabs of SARS-CoV-2 infected SOT showed higher viral loads and longer clearance compared to the general population^4^. The analysis of SARS-CoV-2 specific humoral and cellular responses in SARS-CoV2 infected SOT is only limited to small case series, these data show an intact ability to induce Ab responses but more pronounced weaning over time^5^.

In line with this, it is crucial to define an effective immunization schedule in this population. Whereas the administration of a 2-dose SARS-CoV-2 mRNA vaccination proved to be safe and effective in immunocompetent individuals^6,7^, it may fail in immune compromised patients. After the COVID19 vaccination campaign started in December 2020, vulnerable populations including elderly and immune compromised patients, despite their suboptimal immune responses upon vaccine preventable diseases^8,9^ were prioritized in the national immunization programs. However, preliminary data on solid organ transplant recipients (SOT) showed a lower proportion of detectable SARS-CoV-2 antibodies (Ab) after both 1^st^ and second dose of mRNA vaccine^10,11^. The impact of this suboptimal response as well as their ability to induce a CD4+ Antigen specific response remain unknown.

In the present study we first aimed to confirm the safety of a 2-dose SARS-CoV-2 mRNA vaccination in heart and lung transplanted patients. We further investigated SARS-CoV-2 specific antibodies and frequency of antigen specific CD4 T cells at baseline, 21 days after priming and and 7 days after booster dose. Additional markers of suboptimal immune response upon vaccination, including the impact of distinct immune suppressive regimens and phenotype characteristics of T and B cells were evaluated.

## METHODS

### Study participants

Thirty-four SOT (heart (n=30), heart-kidney (n=2), lung (n=2) were enrolled between February 19^th^ to July29^th^ 2021 at Bambino Gesù Children’s Hospital. Clinical characteristics and baseline laboratory analysis are shown in Table 1. In the present study longitudinal blood samples were collected at the time of the first dose (T0), at the time of the booster dose, 21 days after first dose (T21), 7 days after booster dose (T28). SARS-CoV-2 serology was further tested at a follow up visit scheduled between 60 and 120 days after the priming (T120). All patients were naïve to SARS-CoV-2 infections as demonstrated by the absence of SARS-CoV-2 N-antibodies and no history of COVID19; they all received BNT162b2 mRNA COVID-19 Vaccine, with a schedule of 2 doses of 30mg 21 days apart^12^. Longitudinal blood samples were collected the day of vaccination (T0), 21 days after the first dose (T21) and 7 days after the second dose (T28). An additional serologic test at approximately 120 days (T120) after priming was performed in order to investigate mid-term maintenance of Ab response upon SARS-CoV-2 after vaccination. A negative serology test and molecular tests through nasopharyngeal swabs for SARS-CoV-2 were performed before the vaccination. Health care workers who received BNT162b2 mRNA COVID-19 vaccine were used as control group (HC). All participants received a questionnaire about the Adverse Events and Side Effects following each dose of vaccine. All procedures performed in the study were in accordance with the ethical standards of the institutional research committee and with the 1964 Helsinki declaration and its later amendments. Local ethical committee approved the study and written informed consent was obtained from all participants or legal guardians. Age, gender, clinical and routine laboratory characteristics of the cohort are described in Table 1.

**Table 1.** Patients’ characteristics.

|  | **SOT**<br>**n= 34** | **HC**<br>**n= 36** | **P**<br>**values** |
| --- | --- | --- | --- |
| **Females, n (%)** | 16 (47,1%) | 23<br>(63,9%) | n.s. |
| **Age, mean (+/- SD)** | 29,03 (+/- 7,4) | 47,4 (+/- 12,2) | 0,0001 |
| **Age at transplantation, mean (+/- SD)** | 14,65 (+/- 7,24) | n.a. | n.a. |
| **Age from transplant at vaccination, mean (+/- SD)** | 14,06 (+/- 8,2) | n.a. | n.a. |
| **Type of SOT, n (%)** | Heart, 30 (88,2%)<br>Heart-kidney, 2 (5,8%)<br>Lungs, 2 (5,8%) | n.a. | n.a. |
| **Immune suppressive treatments, n (%)** | Cyclosporine, 27 (79,40%)<br>Everolimus, 26 (76,40%)<br>Mycophenolate, 10 (29,40%)<br>Tacrolimus, 6 (17,64%)<br>Azathioprine, 2 (5,88%)<br>Prednisone, 11 (32,35%) | n.a. | n.a. |
| **Immune suppressive treatment baseline serologic levels**<br>• Cyclosporine ng/mL, mean (+/-SD)<br>• Everolimus ng/mL, mean (+/-SD) | 95,39 (+/- 36,41)<br>3,56 (+/- 1,53) | n.a. | n.a. |
| **Diagnosis, n (%)** | Congenital heart disease, 15 (44,1%)<br>Dilated cardiomyopathy, 15 (35,3%)<br>Restrictive cardiomyopathy, 3 (8,8%)<br>Obstructive hypertrophic cardiomyopathy, 2 (5,9)<br>Cystic fibrosis, 2 (5,9%) | n.a. | n.a. |
| **White Blood Cells /mL, mean (+/- SD)** | 6150, (+/- 1588) | n.a. | n.a. |
| **Hemoglobin mg/dL, mean (+/- SD)** | 13,17 (+/- 1,85) | n.a. | n.a. |
| **Lymphocytes cells/mL, mean (+/- SD)** | 1745 (+/- 892) | n.a. | n.a. |
| • CD 3 %, mean (+/- SD)<br>• CD 4 %, mean (+/- SD)<br>• CD 8 %, mean (+/- SD)<br>• CD 19%, mean (+/- SD)<br>• CD 16+56+, mean (+/- SD) | 72,16 (+/-16,83)<br>39,76 (+/- 10,67)<br>30,26 (+/- 9,11)<br>11,28 (+/- 5,47)<br>13,78 (+/- 8,32) | n.a. | n.a. |

### Safety

Specific local or systemic adverse events and use of antipyretic or pain medication within 7 days after the receipt of each dose of vaccine or placebo were collected. Information were collected through a paper questionnaire reporting both solicited local and systemic adverse Events and Side Effects following each dose of vaccine, up to 7 days after 2nd dose. Events of graft rejection were monitored during follow up visits up to 4 months after vaccination.

### Sample collection and storage

Venous blood was collected in EDTA tubes and processed within 2 hours. Plasma was isolated from blood and stored at -80°C. Peripheral blood mononuclear cells (PBMCs) were isolated from blood of all patients with Ficoll density gradient and cryopreserved in FBS 10% DMSO until analysis, in liquid nitrogen.

### Humoral response

Anti-SARS Cov-2 IgG Antibodies (Ab) titres were measures as previously described^13^ at T0, T21 and T28 and during the follow up visits at approximetly 120 days after the 1^st^ dose (T120). In particular, we measured Ab against the S1-receptor-binding-domain (RBD) (Roche, cut-off: 0.8 U/mL) and anti-trimeric SARS-CoV-2 Ab (LIASION® SARS-C0V-2 DiaSorin, cut-off: 13 AU/mL).

### CD4 Ag-specific T cells by flow cytometry

SARS-CoV-2 specific CD40L+CD4+ T cells were identified, as previously described^14^. Briefly, thawed PBMC were plated (1.5×10^6^/aliquot/200 ul) in 96 well-plate containing CD154-PE (CD40L, BD PharMingen, Franklin Lakes,NJ, USA), anti-CD28 (1mg/ml) and either 0,4mg/ml PepTivator SARS-CoV-2 Prot_S (Milteny Biotech, Ber-gisch Gladbach, Germany) or media only. Following 16hrs incubation at 37°C/5% CO2, PBMC were centrifuged and stained with LIVE/DEAD fixable NEAR-IR dead cell stain kit (for 633 or 635 nm excitation, ThermoFisher, Waltham, Massachusetts, US) 1ul per 10^6^cells/ml for 15 minutes at room temperature (RT), protected by light. Surface staining was performed using the following antibodies: CD3 PE-CF594 (clone UCHT1, BD (562280)), CD4 APC-Cy7 (clone RPA-T4, BD (557871)), CD27 FITC (clone M-T271, BD (555440)), CD45RO PE-Cy5 (clone UCHL1, BD), CD185 BV605 (CXCR5, clone RF8B2, BS), CD10 BV510 (clone HI10a, BD 563032), CD19 APC-R700 (clone SJ25C1, BD 659121), CD21 APC (clone B-Ly4, BD 559867), IgD BV421 (clone IA6-2, BD 565940). Gating strategy for T and B cells population and SARS-CoV-2 specific CD40L+CD4+ T cells were gated as reported^14^.

## QUANTIFICATION AND STATISTICAL ANALYSIS

Statistical analyses were performed using GraphPad Prism 8 (GraphPad Software, Inc., San Diego, CA). Statistical significance was set at p < 0.05 and the tests were two-tailed. All data were analysed by D’Agostino-Pearson to assess normality and respectively parametric and non parametric tests were used for normal or not normally distributed datasets. As indicated in figure legends, paired and non-paired test was used to assess differences between Ab load at the different time points, and between SOT and HC respectively. Graphpad Prism 8 software was used for statistical analysis of the cells type distribution and serological parameters, for demographic and routine laboratory blood tests.

## RESULTS

### Patients’ characteristics

Among the SOT, the mean age at transplantation was 14.65 (SD +/- 7.24), and included 30 heart transplanted patients, 2 kidney-heart and 2 lung SOT recipients. The diagnosis that led to transplantation was congenital heart disease (44.1%), dilative cardiomyopathy (35.3%), obstructive hypertrophic cardiomyopathy (5.9%), chronic renal failure (5.9%) and cystic fibrosis (5.9%). As confirmed by baseline serology and clinical records, none of the patients reported past history of COVID19 disease. At every follow up visit (every 2-3 months) during the course of the pandemics, all patients performed nasopharyngeal swab (NPS) screening. All HC, as healthcare workers, performed a periodic NPS and serology screening over the course of the pandemics. Only one SOT (VAX003) out of 34 had a SARS-CoV-2 infection diagnosed 13 days after 2^nd^ dose. Diagnosis was made during a screening performed for a close familiar contact resulted positive. The patient remained fully asymptomatic. For patient VAX003, anti SARS-CoV2 Ab titers (anti S Ab) was absent at T0, 1.5 AU/mL at T21 and 191AU/m at T120. No SOT had signs or history of graft rejection over a 4 months of follow up. As showed in Table 1, age at vaccination resulted significantly higher in HC (p= 0.0001). All patients were under immune suppressive treatments at the moment of immunization, full list of immune suppressive regimens is reported in Table 1.

### Safety of 2 doses of BNT162B2 MRNA COVID-19 VACCINE

Among the SOT cohort, mild-to-moderate pain at the injection site within 7 days after the first dose was reported in 70% of the patients and the most commonly reported solicited local reaction (Fig 1). A noticeably lower percentage of participants reported injection-site redness (8,6%). The proportion of participants reporting local reactions slightly increased after the second dose (94%), with pain reported in all patients and redness or swelling reported in the 23% of patients. Only 6% of participants reported severe pain after both 1st and 2nd dose. In general, solicited local reactions were mostly mild-to-moderate in severity and resolved within 1 to 3 days. Systemic adverse events were reported in 48.8% of patients after the 1st dose and in 72.2% after the 2nd dose. The majority of patients reported mild to moderate systemic reactions, however the percentage of severe systemic reactions increased after the 2nd dose (11%). The most commonly reported systemic reactions was headache and myalgia, described in 69% of patients after the 2nd dose. Only the 9% and the 23% of patients respectively after the 1st and the second dose reported the self-administration of anti-inflammatory treatments. Full reports of symptoms are described in Table 2. No patients were hospitalized due to adverse reactions. No patients suffered from graft rejection up to four months after vaccination.

**Table 2.** Adverse events description in the SOT cohort.

|  | **N (31) 1 dose** | **N (18) 2 dose** |
| --- | --- | --- |
| **Systemic adverse events\*** | **15 (48,4%)** | **13 (72,2%)** |
| Fever | 1 | 1 |
| Cold-like symptoms | 7 | 5 |
| Asthenia | 7 | 7 |
| Myalgia | 9 | 9 |
| Headache | 6 | 9 |
| Pharyngodynia | 3 | 1 |
| Vomit | 1 | 2 |
| **Local Adverse Events\*** | **23 (74,2%)** | **17(94,4%)** |
| Pain in the site of injection | 23 | 17 |
| Itch | 2 | 3 |
| Hyperemia | 0 | 0 |
| Edema | 0 | 1 |
| Self administered antiinflammatory or antipyretic treatment | 3 | 4 |

**Figure 1.**
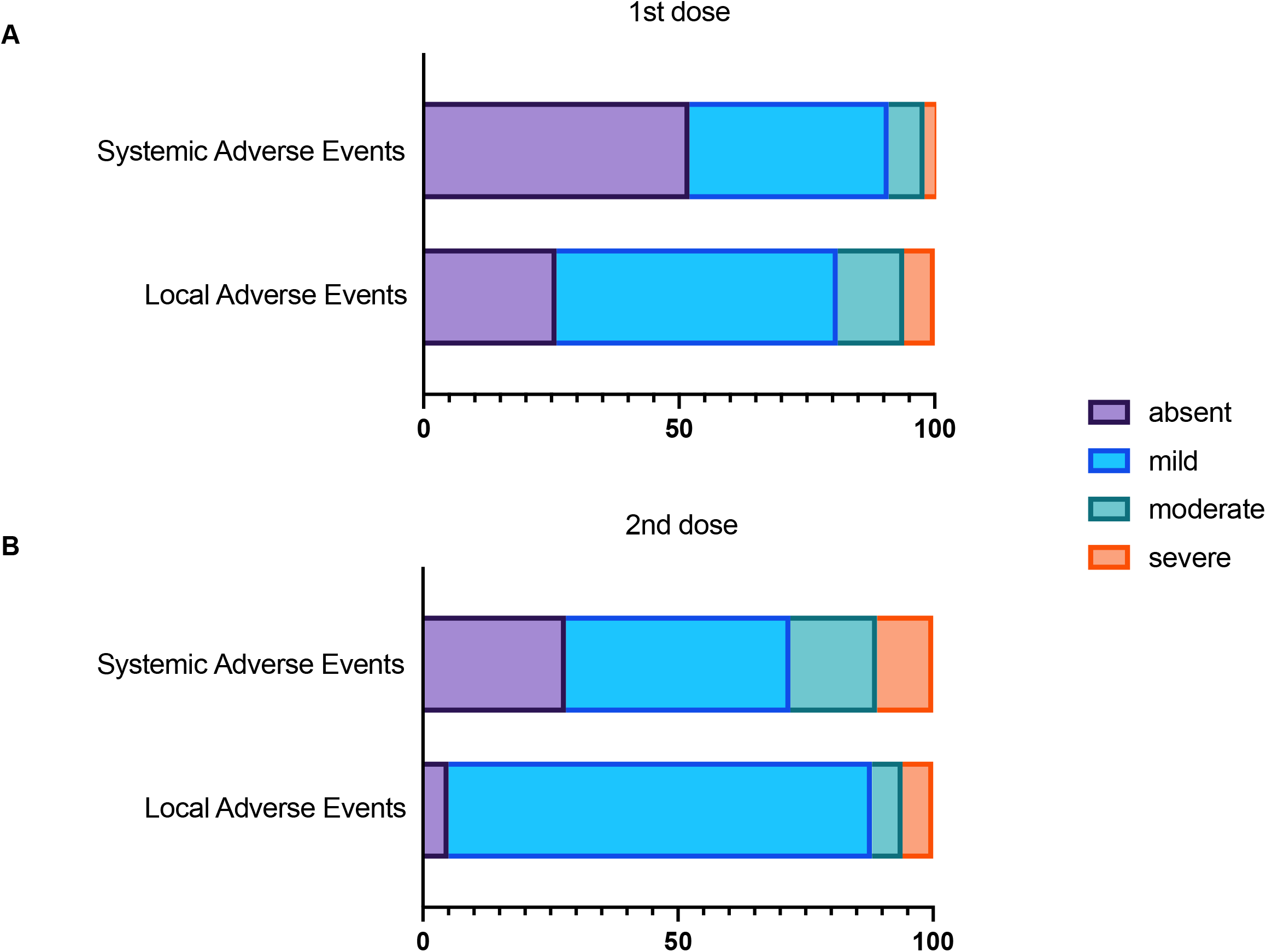
Safety profile of BNT162B2 mRNA COVID-19 vaccine. Contingency plots show safety profile following 1^st^ (A) and 2^nd^ dose (B) of BNT162B2 mRNA COVID-19 VACCINE in SOT divided according systemic or local adverse events.

### Serologic response

At the time of vaccination all patients resulted negative for anti-N, anti-S and anti trimeric antibodies, thus excluding previous immunization or infection upon SARS-CoV-2. Humoral responses after both vaccination doses were overall impaired in SOT compared to HC as summarized in Fig. 2. A significant increase was shown by paired analysis of anti-S in SOT at T21 (p=0.008) albeit to a lesser extent compared to HC (<0.0001) (Fig. 2). In contrast, a significant increase was shown in both SOT and HC in terms of anti-S Ab after 2^nd^ dose of vaccination in both HC and SOT (p<0.0001). Both anti-S and trim Ab titers resulted significantly lower at both T21 and T28 in SOT compared to HC (p<0.0001). All HC showed anti S Ab seroconversion after both 1^st^ and 2^nd^ vaccination dose, whereas only 8 (23.5%) and 22 (68.75%) SOT presented an anti-S Ab titer higher than the limit of detection at T21 and T28 respectively. Ten patients, who showed an anti-S Ab level below the limit of detection at T28, herein referred to as non responder (NR), were further characterized and compared to SOT with detectable Ab response at T28 (responder (R). Fold change anti-S Ab response showed a lower fold change response in both time-points in SOT compared to HC (Fig. 2 G). Further analysis, performed to understand whether a specific immune suppressive regimen was associated to a lower Ab response, showed that SOT treated with immune suppressive regimens including mycophenolate had lower fold change Ab response compared to the rest of SOT (Supplementary Fig. 1). The follow up serologic test, performed in SOT approximately 120 days after vaccination, showed a significant increase in anti S SARS-COV-2 Ab at T120 compared to T28 (p=0.0005). Such response resulted in a reduction of seronegative individuals among SOT, with only 3 out of 32 (9.3%) showing anti S Ab <0.8 AU/mL (Figure 2 A).

**Figure 2.**
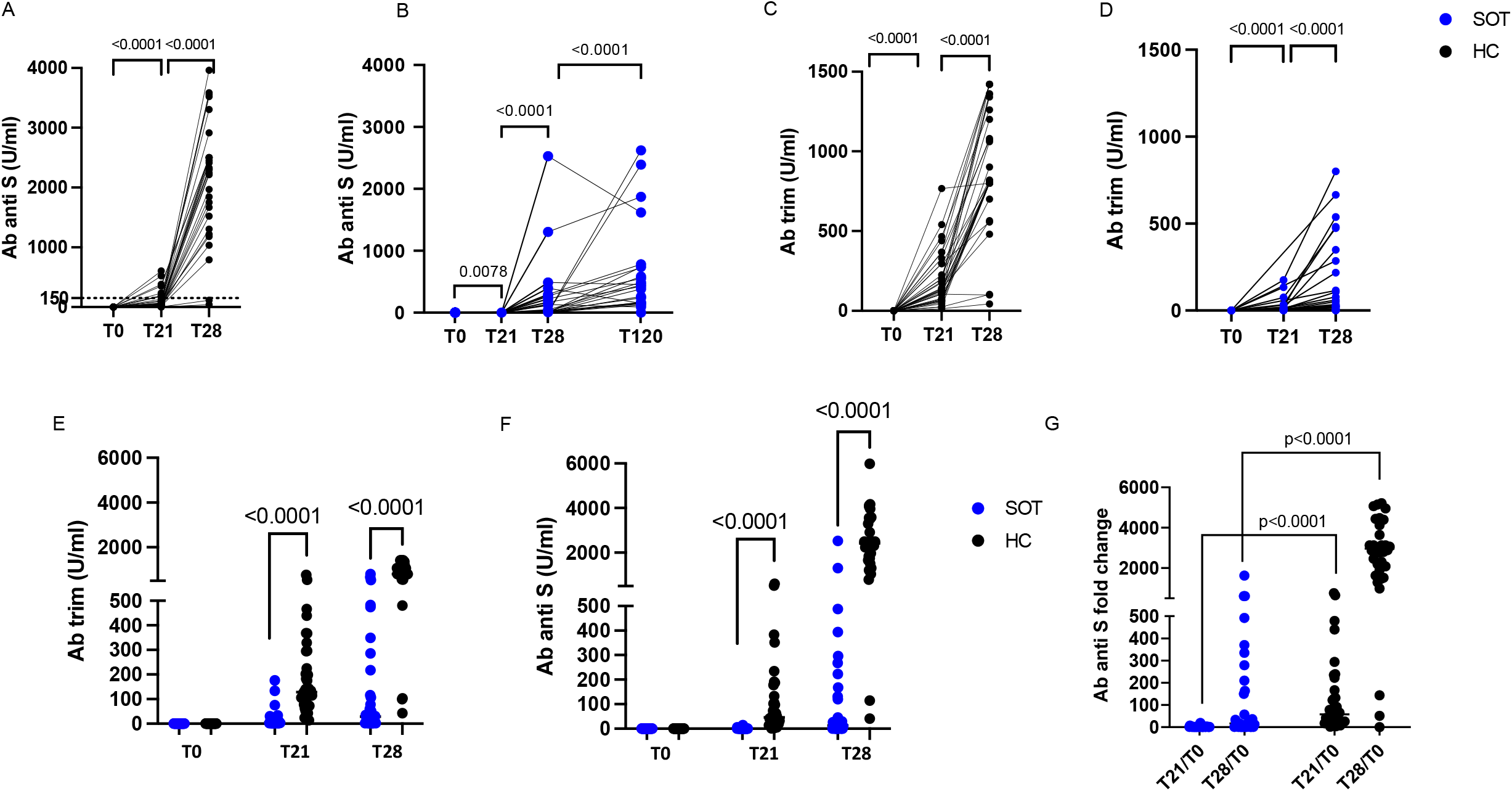
Antibody responses. Longitudinal analysis of anti S (A-B) and Ab anti trim (C-D) respectively in HC and SOT. Paired non parametric T test were performed to define longitudinal Ab increase (A-C). Differences between HC and SOT were calculated for anti trim Ab (E) anti S Ab (F) at T0, T21 and T28. Anti S fold change T21 and T28 with regards to T0 are shown in plot G. Unpaired non parametric T test were used for comparisons.

### Antigen specific T cell immune responses

To further explore the immune response upon the SARS-CoV-2 vaccination we assessed the frequency of SARS-CoV-2 specific T cells responses through the analysis of CD40L positive CD4+ T cells after in vitro stimulation with SARS-CoV-2 peptides, as previously analyzed in infected patients. As previously shown^15^, baseline frequency of CD4+CD40L+ T cells in SOT due to the graft was higher compared to HC (Supplementary Fig. 2). We then assessed the inducible CD40L expression after *in vitro* stimulation in order to detect the frequency of SARS-CoV-2 specific CD4 T cells. Our analysis showed that HC were able to significantly upregulate the CD40L after *in vitro* stimulation with SARS-CoV-2 (p=0.0034), whereas this increase was not observed in SOT (Fig. 3 B). Further analysis were performed on SOT differently responding to the vaccination, however paired analysis showed similar levels of Ag specific T cells between R and NR (Fig. 3 C). Baseline phenotype analysis, performed at the time of priming dose, showed a major imbalance towards memory T cell subsets in SOT compared to HC (Supplementary Fig. 3 A). Indeed, significantly higher levels of central memory, effector memory and lower naïve and terminally differentiated T cells were found in SOT vs HC (Supplementary Fig. 3 B). In line with a chronic external antigen stimulation, fueled by chronic stimulation of the graft, also the CXCR5+ follicular helper CD4 T cells resulted significantly higher in SOT compared to HC (Supplementary Fig. 3 C).

**Figure 3.**
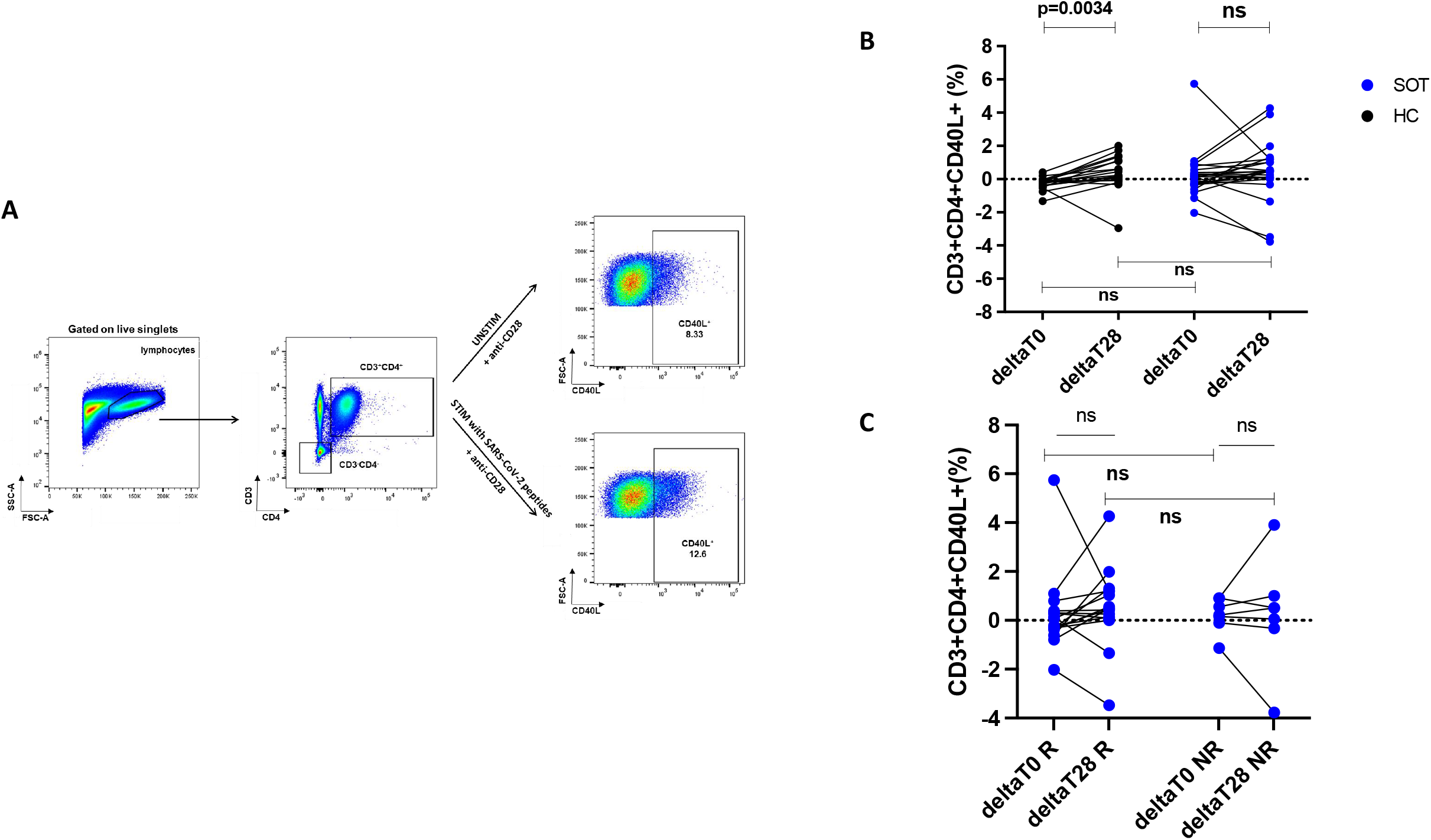
SARS-CoV-2 specific T cell responses. Representative gating strategy for SARS-CoV2 specific T cell is shown in panel A. Paired analysis showing delta of CD40L frequency between stimulated and unstimulated sample is shown in SOT and HC for T0 and T28 (panel B). The same Analysis is shown for SOT not presenting an Ab response (non responders, NR) and for patients with Ab serconversion (responders, R).

### T and B cell phenotype in SOT with distinct response to vaccination

To dissect whether a distinct immune phenotype was able to predict a suboptimal response following the vaccination we performed an extended T and B cell phenotype analysis before vaccination. Our results did not show any difference between NR and R among the different T cell subsets, including total CD3, CD4, CD4 T maturational subsets and T follicular Helper cells (data not shown).

B cell phenotype analysis (representative gate in Fig. 4A) showed similar frequencies of total CD19+ B cells, however the analysis of maturational B cell subsets showed a lower frequency of memory B cell (CD19+CD27+CD21+) in NR compared to R (p=0.008) (Fig. 4 A). This data was also confirmed by a lower frequency of IgD-CD27+ switched memory B cell (p=0.002) (Fig. 4 B).

**Figure 4.**
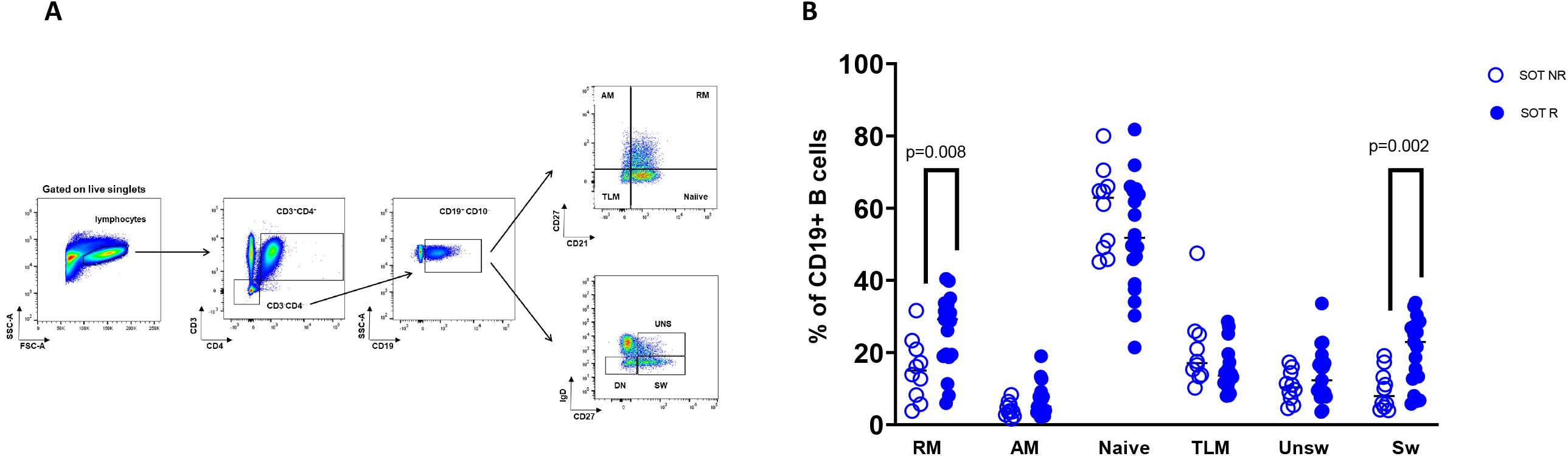
B cell phenotype in SOT with distinct response to SARS-CoV-2 vaccination. Representative gating strategy for B cell subsets is shown in panel A. Dot plot in panel B shows differences in B cell subsets frequencies between SOT NR and R.

## DISCUSSION

In the present study we report the safety profile and the immunogenicity of the **BNT162B2** SARS-CoV2 (Pfizer-Biontech) in a cohort heart and lung transplanted recipients with no previous history of COVID19 up to four months after vaccination. Our data confirmed a good safety profile with no evidence of graft rejection in none of the vaccinated patients as previously showed in a large cohort of adult SOT^16^. Similarly to other studies the anti-S and anti-Trimeric Ab responses were significantly (p<0.0001) lower in SOTs in comparison to HC after both the 1^st^ and the second dose of vaccination. The serologic test performed in a mid-term (2-4 months) follow up visit showed a significant increase in anti S Ab response with only 9% percent of SOT remaining seronegative. These results show a better delayed response compared to a recent study on SOT^17^ where 40% of patients, tested 90 days after the first dose remained seronegative. This difference may be due to the distinct composition of the patients’ cohort with an higher percentage of kidney transplanted, known to undergo more aggressive immune suppressive regimens, and also to the higher percentage of patients being treated with mycophenolate (63% vs 29% in our cohort) and corticosteroids (87% vs 32.3% in our study). In addition to humoral response in this study we firstly evaluate the frequency of SARS-CV2 specific T cell measured trough the upregulation of CD40L. This method, previously validated in infected kids^14^ and in vaccinated healthy controls^13^, has never been tested in SOT and demonstrated lower ability of SOT vs HC to induce Ag specific T cells after in vitro stimulation with SARS-CoV-2 peptides. These data are in line with one previous study which investigated the cellular response after 2 mRNA vaccine doses, and showed that only the 48% of SOT showed a IFN gamma-IL 2 producing CD4 T cells^18^.

Observational studies about vaccine responsiveness against different infectious agents in immunosuppressed individuals may provide insights into further research aiming to tailor vaccine intervention in SOT. Previous studies about influenza vaccine’ safety and immunogenicity in this population represent the major existing backbone of knowledge to investigate COVID19 vaccines. Among these studies a report from 2012-2013 flu vaccination experience overall demonstrated lower immunogenicity in kidney transplanted patients^19^. In a cohort of adult solid-organ transplant recipients a greater seroconversion rates and higher antibody titers in high dose influenza vaccines compared with a standard dose or with an adjuvanted formulation have been reported^20–22^. Such observations highlight the importance to deeply evaluate immunogenicity of the novel Pfizer/Biontech COVID-19 vaccine with alternative strategies, including vaccine dosage. In addition these investigations should also aim at defining potential correlates of effective immune response in this population dissecting both the T-and B-cell lymphocite compartments. As expected, T cell immune phenotype analysis showed a profound impairment in T cell maturational subsets distribution at the time of priming dose, however, such frequencies were not able to distinguish responders from not responders among SOT. On the other hand, a significantly higher frequency of CD27+CD21+ resting memory B cells and CD27+IgD-switched memory B cells was found in responders compared to non-responders patients. Hence a preserved B cell immune phenotype was able to predict vaccine response in SOT similarly to what was previously found in other immune compromised patients with distinct responses upon influenza vaccination^23,24,25^. Additional studies dissecting immune correlates of vaccination failure should be further tested in larger SOT cohorts in order to design tailored vaccination strategies in this population.

With regards to the impact of distinct immune suppressive regimen of SOT on vaccine immunogenicity, our finding which confirmed a lower SARS-COV2 specific immune responses in patients under treatment with antimetabolites such as mycophenolate mofetil deserves discussion^10^. Such correlation doesn’t indicate that these class of drugs should be stopped in order to facilitate a better vaccine response. In fact, withdrawal of antimetabolite therapy may predispose the development of donor specific antibodies, increasing the risk of antibody mediated rejection or cellular rejection of the allograft. Therefore, such clinical actions should only be carried out in well controlled studies under strict clinical monitoring.

Overall our results have confirmed previous studies showing that SOT have a lower response against COVID-19 vaccine. It deserves further discussion what actions can be taken in order to improve such immune response. One could be the use of an adjuvanted vaccine, that should be investigated in these patients. In general, adjuvants used to enhance vaccine immunogenicity also elicit nonspecific inflammatory responses, and thus have the potential to induce acute allograft rejection. Concern about adjuvant safety in organ transplant recipients arose from observational studies where of an unusual increase of anti-HLA antibodies in kidney transplant recipients who received the 2009 influenza A(H1Na1)pdm09 vaccine, which contained the squalene-based AS03 adjuvant system^26,27^. However, only a fraction of these anti-HLA antibodies were donor specific, and a subsequent investigation of >10,000 solid organ transplant recipients found no definitive association between the AS03 adjuvant system and acute allograft rejection. Several recombinant spike protein SARS-CoV-2 vaccines containing adjuvants, such as AS03 are under investigation. Richmond and colleagues recently reported results from a new AS03 adjuvanted vaccine trial: the protein based vaccine alone (no adjuvant) was poorly immunogenic, but neutralising antibodies were recorded after the first injection with the higher doses (9 μg and 30 μg) of AS03-adjuvanted vaccine, which persisted for the remainder of the interim analysis period, with little meaningful difference between younger and older adults (all participants showed seroconversion). Furthermore, the magnitude of neutralising antibody titer and the ratio to binding antibodies was favorable compared with mRNA-based vaccines with reported efficacy^28^. Overall these data suggest that the use of adjuvated vaccine should be investigated for SOTs^29^.

Additional strategies aiming to test the advantage of a “mixed” immunization with a vectored vaccine for priming a and a mRNA vaccine for booster are now being evaluated^30,31^ in heathy individuals and may also be considered in population at risk for lower vaccination efficacy such as SOT.

A number of open questions still need to be elucidated in SOT. Vaccine dosage, duration of vaccine induced immunity, conditions/markers associated with a lower responsiveness as well as vaccination strategies for those who do not properly respond upon COVID19 vaccination are items in search for answers. Furthermore, clinically relevant outcomes of disease severity and mortality need to be assessed for SOT vaccines. One specific aspect to dissect is efficacy of additional booster doses in this population. A recent study showed that a 3^rd^ booster dose of mRNA COVID-19 vaccine, administered 2 months after the 2^nd^ dose was able to induce seroconversion in 45,25% of patients who failed to seroconvert after the standard 2-doses schedule^17,32^. It remains unknown whether this alternative strategy should be directed only to patients with a suboptimal serologic and cellular response, hence suggesting that a periodic longitudinal SARS-CoV-2 immune response surveillance may be needed in vaccinated SOT.

Meantime, in the absence of effective vaccination strategies, consideration may be directed to a valid cocoon immunization strategy or to the prophylactic administration of monoclonal antibodies in selected heavily immunosuppressed patients who experience a close contact with a proven SARS-CoV2 case. The low cellular and humoral immune responses reported in immunosuppressed individuals suggest that patients need to continue to follow□current prevention measures (including wearing a mask, maintaining social distancing and avoiding crowds and poorly ventilated indoor spaces) once vaccinated.

This study presents some limitations mainly due to the small sample size, the short time of follow up and the absence of adequately represented groups of distinct immune suppressive regimens. Larger studies are needed in order to identify a) the proper timing of vaccination to elicit protective immune responses, b) informative correlates of protection and c) alternative immunization strategies for those patients who did not mount protective immune responses upon mRNA vaccine.

## Data Availability

Data used for the submitted paper are available upon request to the Corresponding Author.

## ACKNOLEDGMENTS

We would like to thank all patients and guardians who participated to the study and all the CONVERS study nurses team. We also thank Jennifer Faudella and Giulia Neccia for their administrative assistance.

## CONVERS study Team

Lorenza Romani, MD, Sara Chiurchiu, MD, Andrea Finocchi, MD, PhD, Caterina Cancrini, MD, PhD, Laura Lancella, MD, Maia De Luca, MD, MD, Daniela De Angelis MD, Daniele Selvaggio, Andrea Gioacchino Rotulo MD, Elisa Profeti MD, Enrica Franzese MD, Emanuela Marchettini MD, Ilaria Pepponi PhD, Alessandro Jenkner MD.

## Supplementary Figures

**Supplementary Figure 1**. Anti S Antibody fold change in patients with Immune suppressive regimens including or not including mycophenolate.

**Supplementary Figure 2**. CD40L expression in stimulated aliquots of samples collected at T0 and T28.

**Supplementary Figure 3**. Representative gating strategy of T cell subsets (A). T cell subsets frequencies and T follicular Helper cells (central memory CXCR5+ T cells) are shown respectively in panel B and C.

## ABBREVIATIONS

Anti-HLA: anti human leukocyte antigen
COVID19: coronavirus disease 2019
EDTA: Ethylenediamine tetraacetic acid
HC: healthy controls
IFN: interferon
IL2: interleukin 2
mRNA: messenger Ribonucleic Acid
PBMCs: Peripheral Blood Mononuclear Cells
SARS-CoV-2: Severe Acute Respiratory Syndrome Coronavirus-2
SOT: Solid Organ Transplant Recipients

