## Supplementary figures and images for "HUMORAL AND CELLULAR IMMUNOGENICITY and SAFETY UP TO 4 MONTHS AFTER VACCINATION WITH BNT162B2 mRNA COVID-19 VACCINE IN HEART AND LUNG TRANSPLANTED YOUNG ADULTS"

### Supplemental Figure 1

Supplementary Fig. 1

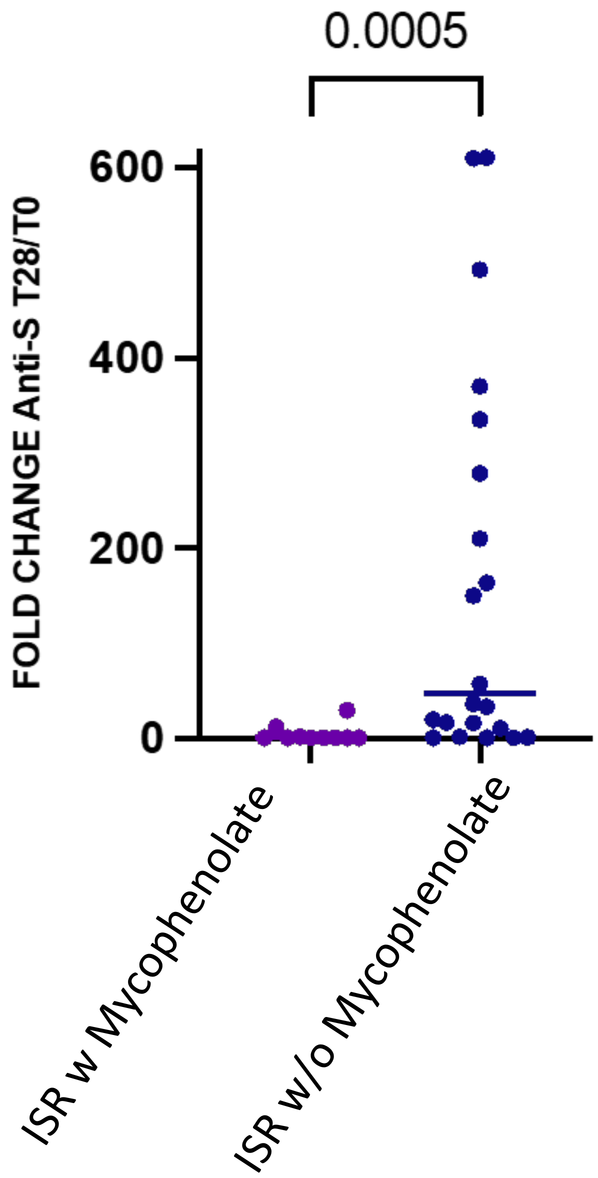

### Supplemental Figure 2

Supplementary Fig. 2

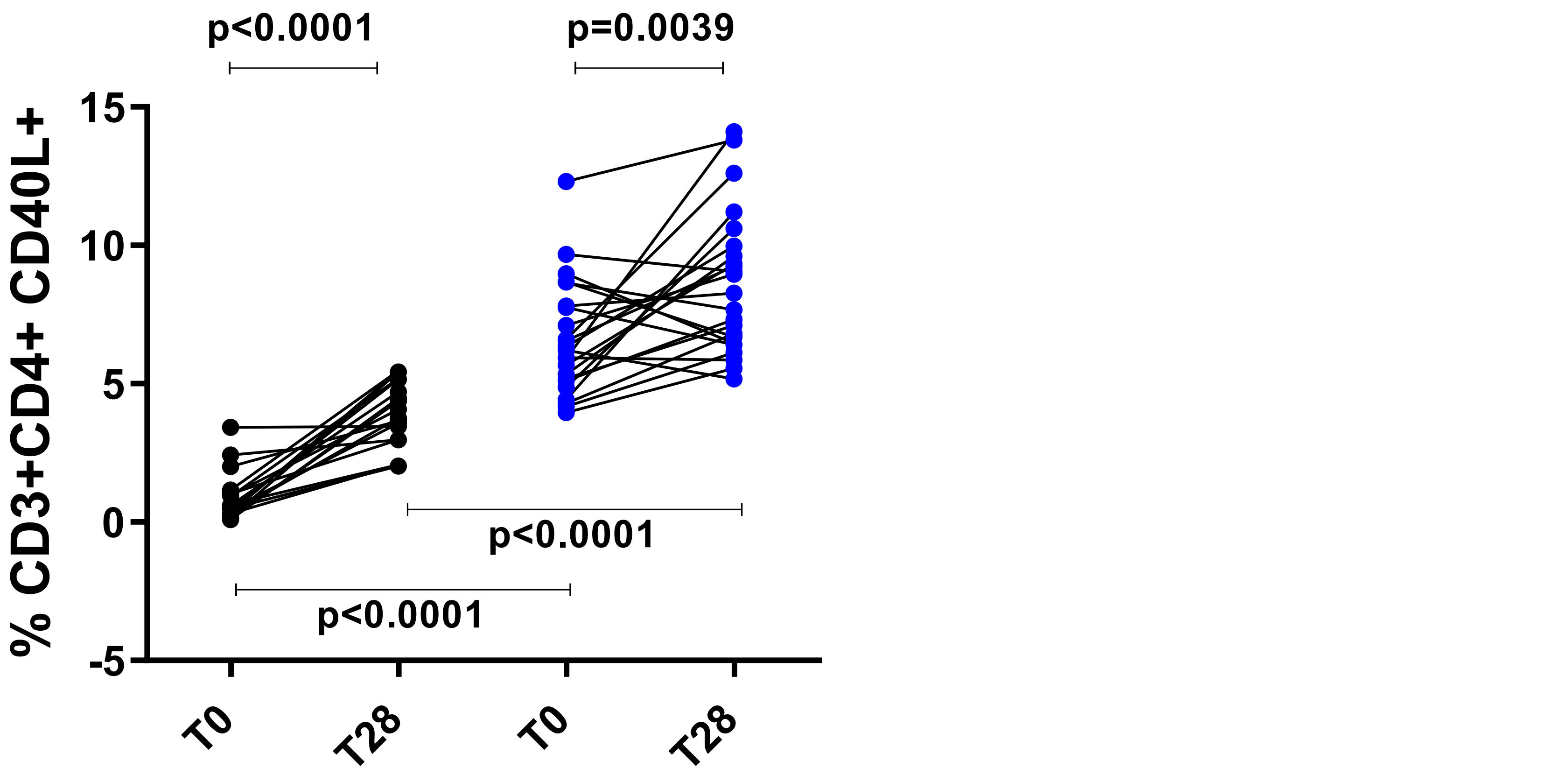

### Supplemental Figure 3

Supplementary Fig. 3

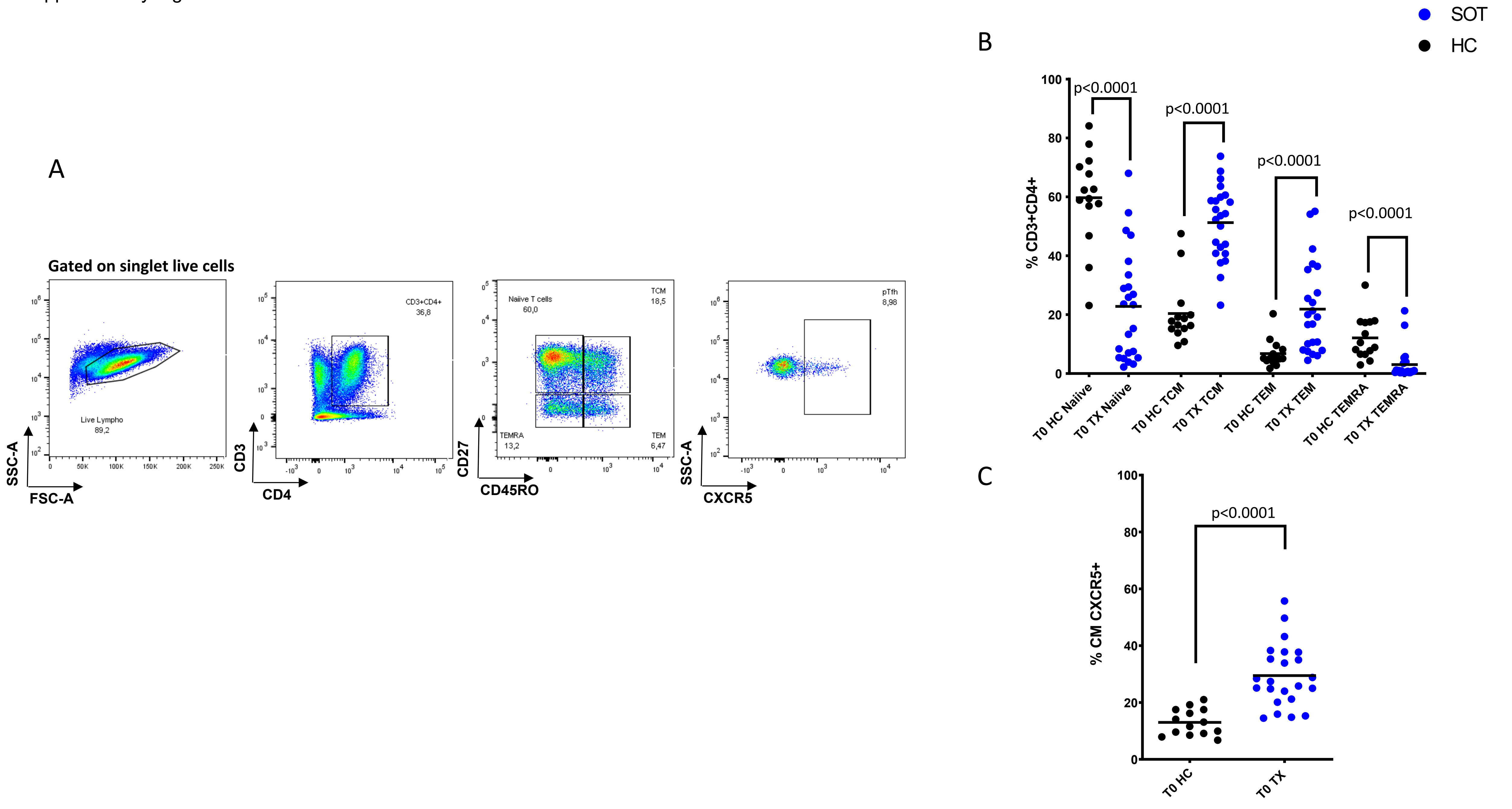
